# Non-invasive tumor probability maps developed using autopsy tissue identify novel areas of tumor beyond the imaging-defined margin

**DOI:** 10.1101/2022.08.17.22278910

**Authors:** Samuel A. Bobholz, Allison K. Lowman, Jennifer M. Connelly, Savannah R. Duenweg, Fitzgerald Kyereme, Aleksandra Winiarz, Margaret A. Stebbins, Biprojit Nath, Michael Brehler, John Bukowy, Elizabeth J. Cochran, Dylan Coss, Janine M. Lupo, Joanna J. Phillips, Benjamin M. Ellingson, Max Krucoff, Wade M. Mueller, Mohit Agarwal, Anjishnu Banerjee, Peter S. LaViolette

## Abstract

**Background:** This study identified a clinically significant subset of glioma patients with tumor outside of contrast-enhancement present at autopsy, and subsequently developed a method for detecting non-enhancing tumor using radio-pathomic mapping. We tested the hypothesis that autopsy-based radio-pathomic tumor probability maps would be able to non-invasively identify areas of infiltrative tumor beyond traditional imaging signatures.

**Methods:** A total of 159 tissue samples from 65 subjects were aligned to MRI acquired nearest to death for this study. Demographic and survival characteristics for patients with and without tumor beyond the contrast-enhancing margin were computed. An ensemble algorithm was used to predict pixelwise tumor presence from pathological annotations using segmented cellularity (Cell), extracellular fluid (ECF), and cytoplasm (Cyt) density as input (6 train/3 test subjects). A second level of ensemble algorithms were used to predict voxel-wise Cell, ECF, and Cyt on the full dataset (43 train/22 test subjects) using 5-by-5 voxel tiles from T1, T1+C, FLAIR, and ADC as input. The models were then combined to generate non-invasive whole brain maps of tumor probability.

**Results:** Tumor outside of contrast was identified in 41.5 percent of patients, who showed worse survival outcomes (HR=3.90, p*<*0.001). Tumor probability maps reliably tracked non-enhancing tumor in the test set, external data collected pre-surgery, and longitudinal data to identify treatment-related changes and anticipate recurrence.

**Conclusions:** This study developed a multi-1 stage model for mapping gliomas using autopsy tissue samples as ground truth, which was able to identify regions of tumor beyond traditional imaging signatures.

## 1. Introduction

Glial brain tumors are the most common primary central nervous system tumors, with an incidence of around 6 per 100,000 persons^1^. High-grade gliomas such as glioblastomas (GBMs) are difficult to treat due to their aggressiveness and pathological heterogeneity, contributing to one- and five-year survival rates of 41 and 5 percent, respectively^2-3^. Current standard of care requires precise tumor localization to maximize efficacy of frontline treatments such as surgical resection and targeted radiation therapy. Studies examining resection extent in gliomas have demonstrated maximizing tumor removal improves patient survival outcomes, underscoring the importance of identifying the full extent of gliomas^4–7^.

MRI is the primary method for non-invasively monitoring gliomas. Gadolinium contrast agents are used to highlight angiogenic disruptions in the blood-brain barrier, which results in an enhanced T1-weighted signal that defines the primary tumor mass^8–10^. Hyperintensities on T2-weighted FLAIR images correspond to a mixture of tumor and edema, though differentiation remains difficult^11–14^. ADC images derived from diffusion-weighted imaging identify areas of diffusion restriction associated with hypercellularity, though studies validating this beyond contrast-enhancing margins have suggested a weaker relationship^14–18^. Machine learning approaches have also sought to maximize the amount of clinically relevant information extracted from non-invasive imaging, employing recent advances in computing to segment radiologist-defined margins, identify patient-level genetic signatures, and predict cellular-level information using biopsies as validation^19–23^.

Despite the promise of these recent techniques, identifying areas of tumor beyond contrast- and FLAIR-hyperintense margins remains difficult. Studies of autopsy samples aligned to MRI acquired near death found areas of active tumor as far as 10 cm beyond the treated margin, indicating need for tumor tracking improvements^16,24,25^. Therefore, studies with access to tissue beyond contrast enhancement are essential for identifying true extent of tumor, particularly in the post-treatment state. This study uses autopsy tissue samples, collected within and beyond the traditional tumor margin, to assess the prevalence of tumor outside the MRI-defined margin and develop a multi-stage model for non-invasive tumor probability mapping. Specifically, we tested the hypothesis that an MRI-based model for tumor probability trained on autopsy tissue samples can track glioma invasion beyond the currently defined margin associated with worse prognoses.

## 2. Methods

### 2.1. Patient Population

This study was approved by the Institutional Review Board of Medical College of Wisconsin. Written, informed consent was obtained from 65 patients to participate in this study as part of our ongoing brain tumor bank starting in 2010, each diagnosed with a primary brain tumor in concordance with the 2021 WHO classification standards for brain tumors.

The study size was selected based on maximizing the number of complete imaging datasets with high quality aligned tissue samples from autopsy included to boost model performance. Clinical and demographic characteristics of this dataset, as well as for relevant subdivisions, are presented in **Supplemental Table 1**. Portions of this study sample have been used to explore radio-pathomic characteristics in prior publications^14,24^, but this is the first study to develop non-invasive maps of tumor pathology using this data. A schematic of the overall study outline and data processing steps is presented in **Figure 1**. Tumor probability mapping software, study data, and other associated code required for preprocessing data will be made available upon request.

**Figure 1:**
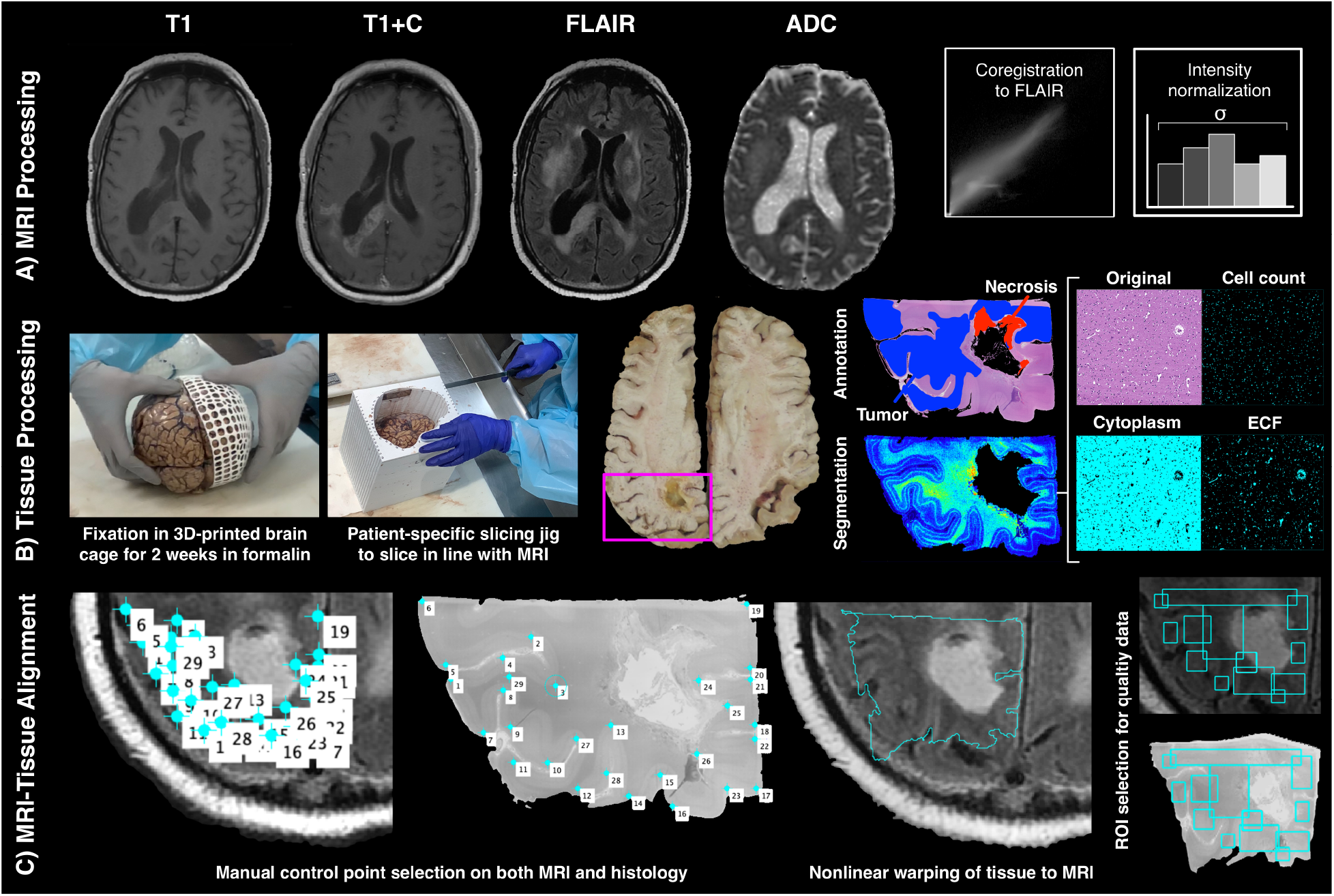
Overview of data collection process. A) T1, T1+C, FLAIR, and ADC scans are collected from each patient’s last imaging session prior to death. All scans are aligned to the FLAIR image and qualitative scans are intensity normalized by the within-brain standard deviation. B) Brains are formalin-fixed in 3D printed brain cages to prevent distortion, and patient specific slicing jigs are used to slice brains in alignment with each patient’s MRI. Tissue is then digitized and segmented into different tissue features, and a subset of patients is annotated for tumor presence. C) Tissue data is then warped to the MRI using manually defined control points, and regions of interest are drawn to selectively include areas of high-quality tissue and MRI data.

### 2.2. MR Image Acquisition and Preprocessing

Clinical imaging was collected from each patient’s last MRI session prior to death. T1, T1+C, FLAIR, and ADC images were selected as the primary focus for this study, as they provide a wide range of information regarding tissue characteristics while remaining ubiquitous across clinical acquisitions. T1, T1+C, and ADC images were each rigidly aligned to the FLAIR image using SPM12 (https://www.fil.ion.ucl.ac.uk/spm/software/spm12/). Qualitative images (T1, T1+C, and FLAIR) were then divided by the intensity standard deviation within the brain to normalize values across patients^8,14^.

### 2.3. Tissue Segmentation and Processing

A total of 159 tissue samples were collected at autopsy using previously published methods^14,16,25^. Board certified pathologists (DC, EJC) collected large-format tissue samples, with additional MRI-based sampling guidance from research staff (SAB, AKL, PSL). Tissue samples were processed, digitized, and segmented using a previously published tool^24^. Additionally, 33 tissue samples from a subset of 9 participants were annotated for presence of infiltrative tumor, tumor with pseudopalisading necrosis (PN), non-tumor necrosis, and unlabeled tissue by a pathologist-trained technician.

### 2.4. MRI-Histology Co-registration

Tissue samples were aligned to each participant’s FLAIR image using previously published Matlab software^14,16,25–27^. Manually defined control points were used to identify architectural landmarks on the tissue data and MRI using autopsy photos to guide accurate placement. Control points were used to compute a non-linear transform that warped tissue to the same space as the MRI, and regions of interest were drawn to exclude areas of tissue distortion (i.e., rips, folds) and MRI artifacts. Following MRI-tissue co-registration, all MRI and tissue data was resampled to 0.4397 × 0.4397 mm per pixel resolution to harmonize all subjects to the most common acquisition parameters.

### 2.5. Assessing prevalence of tumor beyond MRI-defined margin

Aligned autopsy tissue was compared to the T1+C MRI to estimate the prevalence of tumor outside of contrast enhancement (TOC). Tissue samples were inspected for tumor presence using histological characteristics and available immunohistochemical staining (i.e., Ki-67, CD31). Patients were coded for presence of TOC by comparing manually segmented maps of contrast to aligned tissue samples. Chi-squared tests were used to determine differences in TOC frequency amongst patients who did and did not receive radiation with temozolomide (Rad+TMZ), Bevacizumab (Bev), and tumor treating fields (TTFields), as well as amongst diagnostic grade groups. A Cox proportional hazards regression was fit to compare survival durations between patients with and without TOC, including time between MRI and death as a covariate. Survival curves were limited to patients who received standard treatment (Rad+TMZ) to exclude a small number of untreated patients who survived shorter than similarly treated patients. Analysis was performed for the full data set (N=65), as well as on subjects with scans less than 90 days before death (N=47) and patients with a primary GBM at first surgery (N=37) to compare results across potential confounds.

### 2.6. Predicting tumor presence from pathological segmentations

A chart indicating the flow of patients across the multiple model training stages is presented in **Supplemental Figure 1**. The first component of the multi-stage tumor prediction model involved predicting tumor annotations from the segmented pathology data. Several different candidate models were assessed, including k-nearest neighbors, naïve bayes, decision trees, and random-under-sampling-boosted random forest (RUS Tree) models. Each model was trained using pixelwise cellularity, cytoplasm density (Cyt), and extracellular fluid density (ECF) as input to predict tumor (infiltrative tumor/PN) versus non-tumor (non-tumor necrosis/unlabeled). Models were trained on slides from 6 subjects and validated on the 3 remaining subjects. Model performance was evaluated using receiver operator characteristic (ROC) plots and area under the curve (AUC) metrics. The model with the highest AUC was then incorporated into the multi-stage model.

### 2.7. Predicting pathological segmentations from MRI data

The second stage of the multi-stage model focused on training separate models to predict cell, Cyt, and ECF density using MRI data. Bootstrap aggregating random forest models were trained to predict voxel-wise densities using 5-by-5 voxel tiles from T1, T1+C, FLAIR and ADC as input. This framework was selected based on our previous publication, which developed a proof-of-concept for predicting cellularity using MRI data in a smaller patient sample^24^. These models were developed on 2/3rds of the full dataset (N=43) and tested on the held-out set (n=22) to assess generalizability. No patients in the pathological tumor prediction test set overlapped with the training set for these radio-pathomic models, ensuring the test set contained only data unseen in training for every component model. Quantitative performance was evaluated using root-mean-squared error (RMSE) estimates within each test set subject, and example segmentations were plotted and compared to ground truth segmentations.

### 2.8. Predicting tumor presence from MRI data

Following model training, the best performing pathological tumor prediction model was used to convert whole brain cell, ECF, and Cyt density maps to tumor probability maps (TPMs). These maps were plotted against ground truth tissue segmentations and annotations to assess accuracy at identifying novel tumor areas. Additionally, TPMs were generated for three external subjects to assess the quality of the maps and ability to identify non-enhancing tumor on MRI data acquired at other institutions. The first external subject was a 61-year-old male diagnosed with a recurrent GBM, whose scan was collected by our external collaborators at the University of California - Los Angeles. The second external case was a pre-treatment acquisition from a 50-year-old male diagnosed with GBM, taken from the publicly available TCGA-GBM dataset (https://cancergenome.nih.gov/)^28,29^. The third case was a 53-year-old female diagnosed with an IDH1-wildtype GBM, whose scan was collected by an external collaborator from University of California - San Francisco, for which biopsy data from surgical resection was available for an area beyond contrast enhancement. Lastly, TPMs were generated from an independent local dataset of 84 GBM cases^30^ with imaging acquired pre-surgery to determine areas of predicted tumor outside of contrast enhancement (pTOC). A Kaplan-Meier curve was fit to assess differences in overall survival between patients with and without pTOC (n=58 and 26, respectively).

## 3. Results

### 3.1. Prevalence of non enhancing tumor

**Figure 2** shows the prevalence estimates for TOC presence, as well as corresponding Kaplan-Meier plots for survival analyses. TOC was observed in 41.5 percent of patients, with increased presence amongst GBM (GIV) patients (χ^2^=10.73, p=0.005), patients who have been treated with Rad+TMZ (χ^2^=3.99, p=0.046), and patients treated with bevacizumab (Bev) (χ^2^=5.41, p=0.020). Treated patients with TOC showed decreased survival rates compared to patients without TOC (HR=3.90, 95 percent CI = 2.50-5.30, p*<*0.001). Patients with MRI data from within 90 days of death also showed increased TOC presence amongst Rad-TMZ and bevacizumab-treated (χ^2^=4.50, p=0.033 and χ^2^=4.50, p=0.033, respectively) patients, and reduced survival associated with TOC (HR = 3.85, 95 percent CI = 2.33-5.37, p = 0.001). Primary GBM patients did not show significant differences in TOC presence with respect to treatment but did show reduced survival within patients with TOC (HR = 2.95, 95 percent CI = 1.32-4.57, p = 0.025).

**Figure 2:**
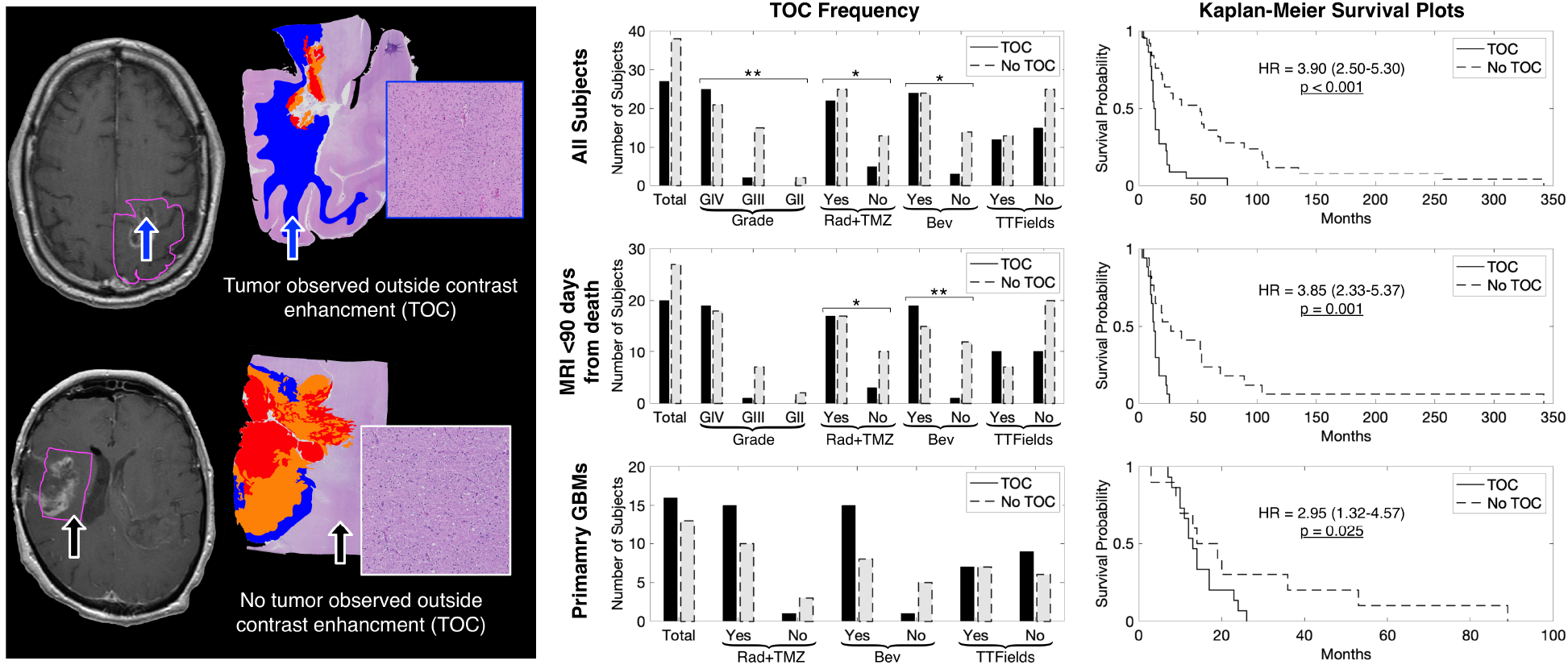
Prevalence and survival analyses for patients with tumor outside contrast enhancement. Analyses were performed separately for the full dataset, patients with MRI less than 90 days prior to death, and patients with a primary GBM. Survival analyses were conducted using Cox proportional hazards regression and only include patients who have received Rad+TMZ treatment. Results indicate increased TOC frequency amongst GBMs and patients who have received Rad+TMZ or Bev treatment. Patients with TOC also show reduced survival compared to patients without TOC. *p*<*0.05, **p*<*0.01

### 3.2. Predicting tumor presence from pathological segmentations

**Figure 3** shows the performance results for the pathological tumor prediction model, along with example tumor probability maps and corresponding ground truth annotations. The RUS Tree model gave the best ROC AUC of 0.857 and was therefore selected as the model used to convert MRI-based pathological segmentations into tumor probability maps. Example predictions show good concordance between areas of high tumor probability and actual tumor presence within both IT and PN tumor types, while correctly avoiding edema and other non-tumor areas.

**Figure 3:**
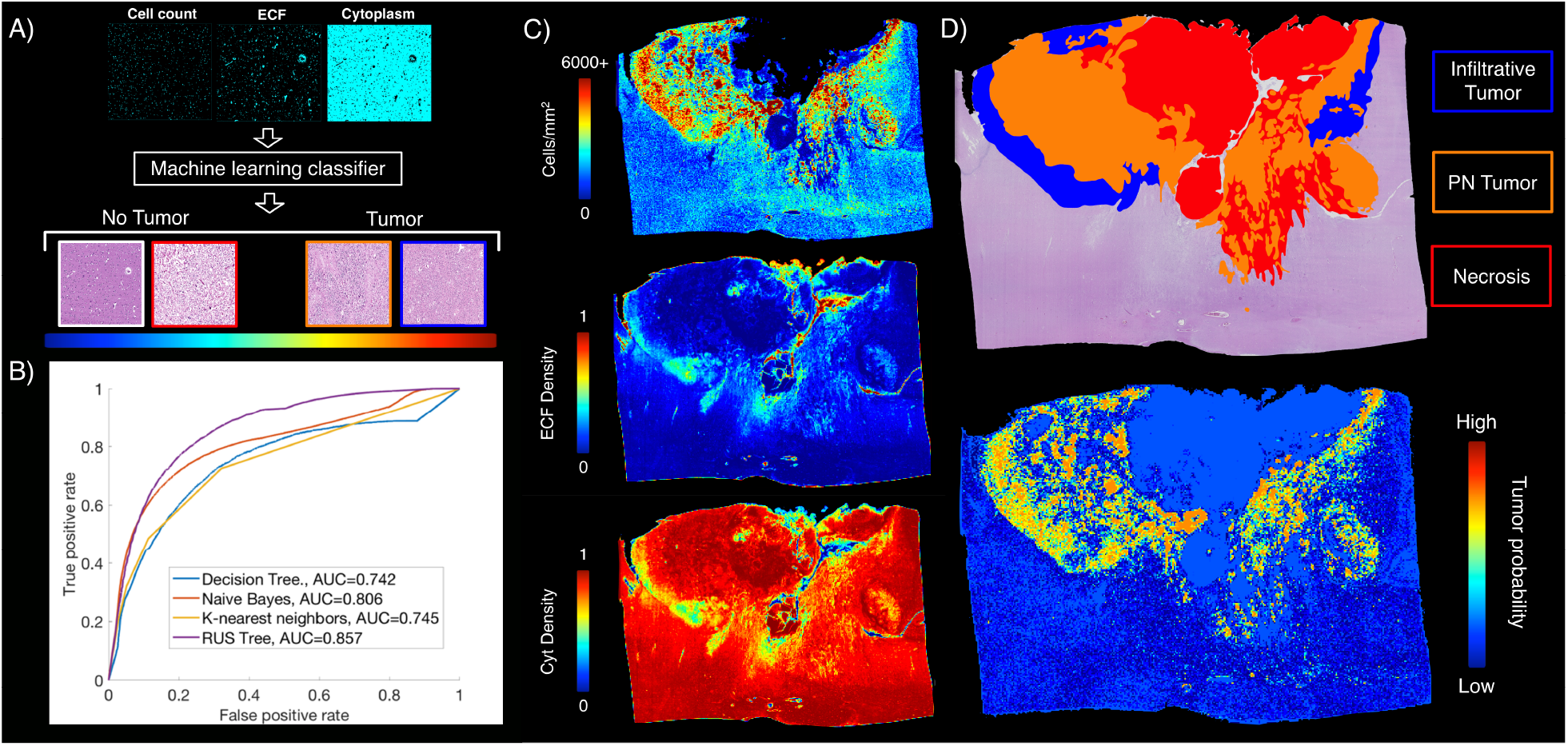
Pathological tumor prediction. A) The pathological tumor prediction model uses cell density, ECF, and Cyt segmentations to distinguish tumor vs. non-tumor using the pathological annotations as ground truth. B) The RUS Tree algorithm was the highest performing tumor prediction model (AUC=0.857) and was used in the final multi-stage TPM model. C) Example tumor probability maps from the RUS Tree model show accurate tumor prediction in both IT and PN areas, while avoiding both normal tissue and areas of necrosis.

### 3.3. Predicting pathological segmentations from MRI data

Segmentation prediction results are shown in **Figure 4**, along with example whole brain predictions on test set subjects. Each radio-pathomic model had a mean subject-level RMSE value within a standard deviation of the ground truth (cell density Std. RMSE = 0.756, Cyt Std. RMSE = 0.917, ECF Std. RMSE = 0.941), indicating satisfactory model performance for each tissue type. Example predictions indicate regions where these maps improve pathological interpretations of the MRI data, such as discrimination between areas of edema and hypercellularity within the FLAIR hyperintense region and areas of hypercellularity beyond contrast enhancement.

**Figure 4:**
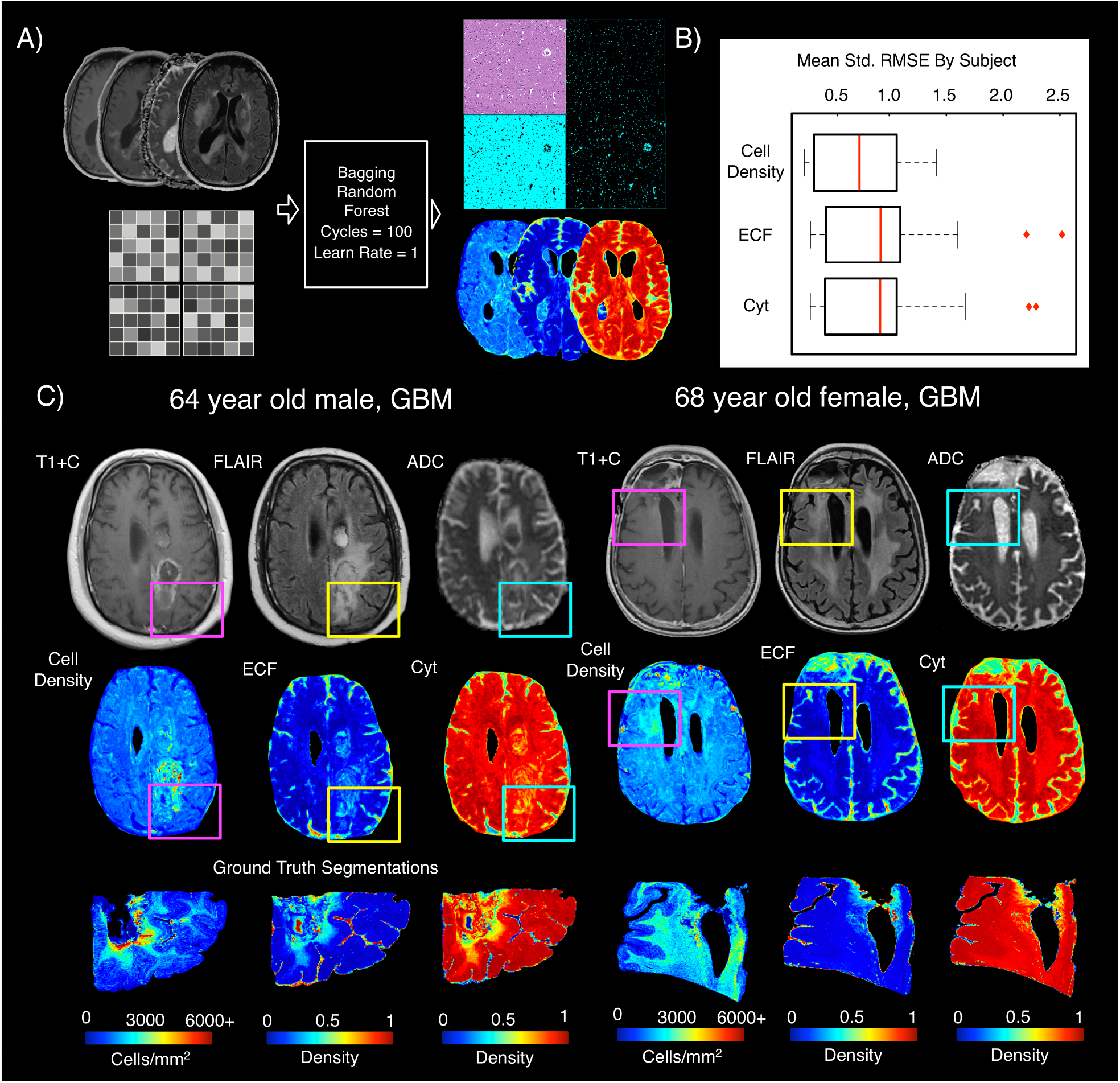
Radio-pathomic maps of tissue segmentations. A) 5 by 5 voxel tiles from T1, T1+C, FLAIR, and ADC were used to predict voxelwise cell density, ECF, and Cyt using bagging random forests. B) Test set performance indicates an average subject-level RMSE within a standard deviation of the tissue ground truth for each tissue class, indicating satisfactory model performance for most subjects. C) Example tissue predictions show areas of accurately predicted cellularity beyond contrast enhancement, as well as distinguishing between vasogenic edema and hypercellular areas within the FLAIR hyperintense region. The subject on the left shows a portion of hypercellularity extending posterior to the contrast-enhancing margin, while highlighting a reduction in cell density within the FLAIR hyperintensity anterior to the contrastenhancement. The subject on the right shows an area of increased cellularity in the absence of contrast enhancement and diffusion restriction.

### 3.4. Predicting tumor presence from MRI data

An example TPM generated from the full multi-stage model are presented in **Figure 5**. Test set TPM with pathological annotations available showed good correspondence between areas of predicted and actual tumor, with separation between areas of PN (high cellularity, high ECF) and areas of non-PN tumor (high cellularity, normal ECF) seen on the corresponding segmentation maps. Tumor was also accurately observed beyond the contrast enhancing region. External data and longitudinal TPMs are presented in **Figure 6**. Predictions on external data sets demonstrated that TPMs generated from this framework provided low-noise, interpretable maps on new data, as well as identified new areas of possible tumor infiltration beyond contrast enhancement. Longitudinal predictions for the example subject showed the TPM in response to treatment, where areas of heightened tumor probability tended to fade after administration of Bev+TMZ and returned following treatment cessation. These maps also identified tumor crossing-over into the contralateral hemisphere as early as 374 days prior to death despite a lack of contralateral contrast enhancement across the patient’s imaging. This area of crossing over was confirmed to be tumor at autopsy.

**Figure 5:**
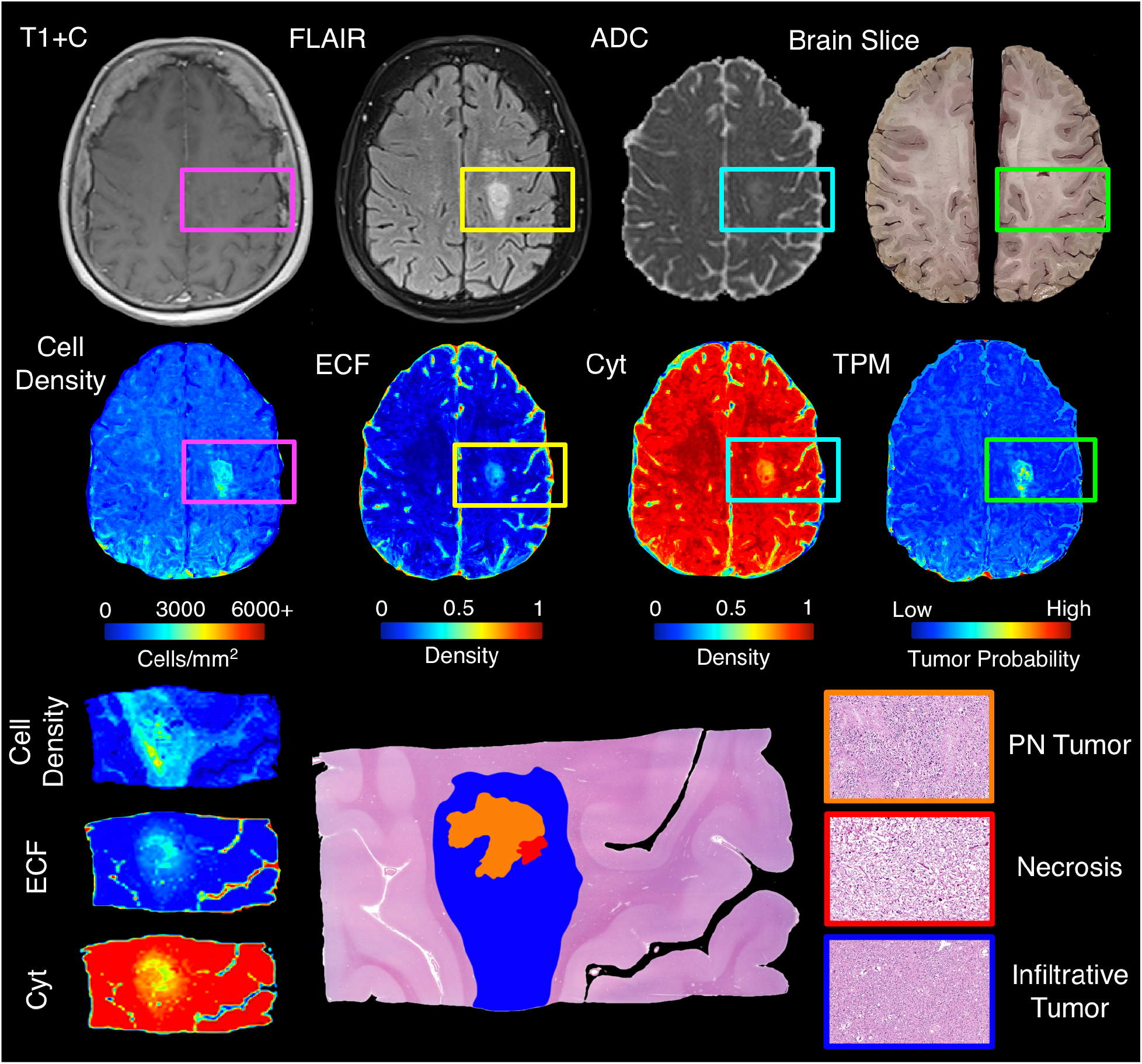
Example TPM for a test set subject (GBM, Female, 80yo), along with corresponding MRI and segmentation predictions. The TPM for this patient accurately highlights an area of tumor outside contrast enhancement and in the absence of diffusion restriction. In addition, the cell density map highlights that the entire tumor area is hypercellular, while the ECF map highlights a high-ECF core to the hypercellular area, which correctly suggests an area of pseudopalisading necrosis (high cellularity, high ECF). This demonstrates the ability for TPMs to distinguish between pathologically distinct regions of tumor.

**Figure 6:**
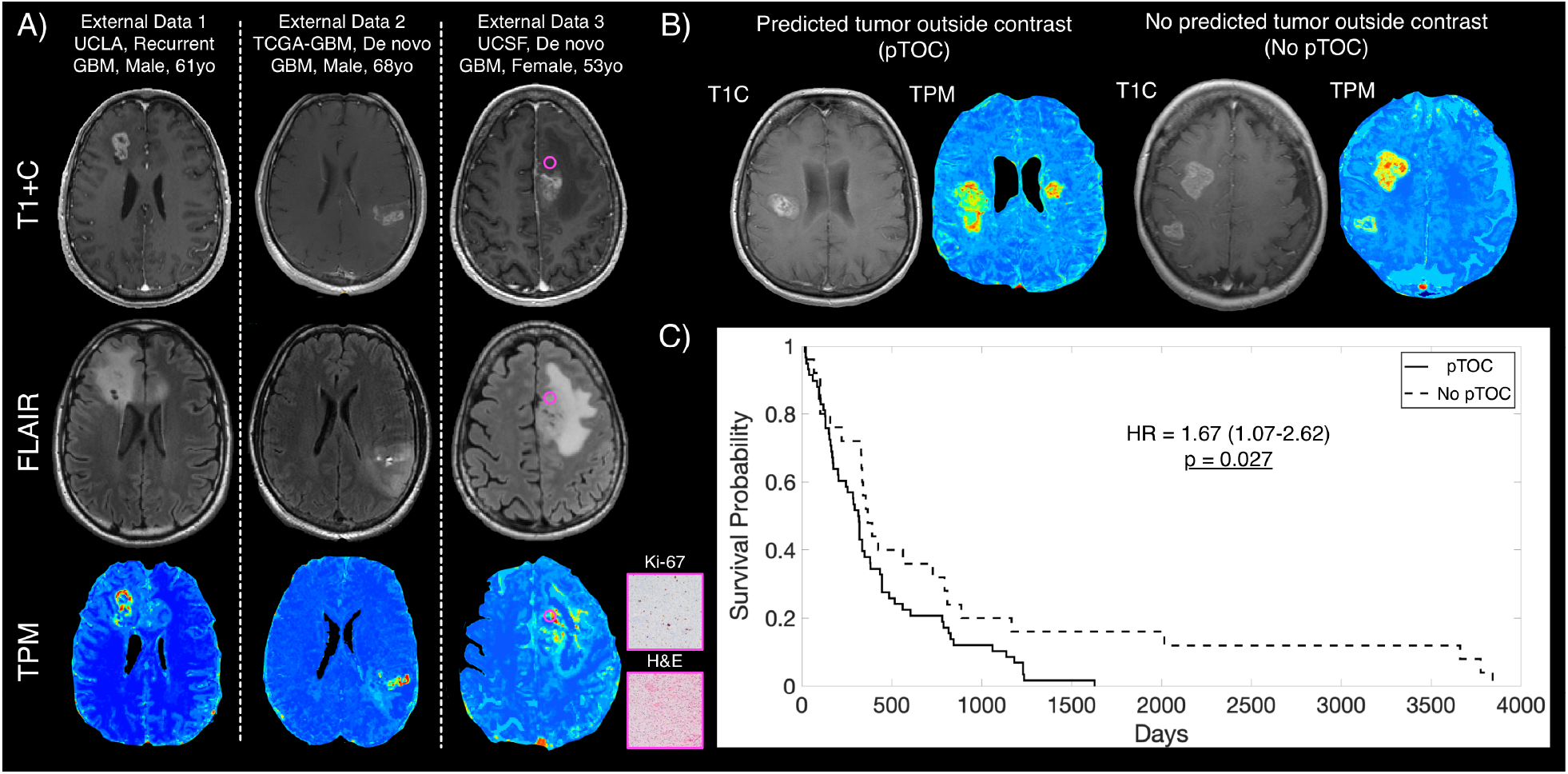
External TPMs and survival analyses for predicted tumor outside of contrast enhancement pre-surgery. A) Example TPMs applied to externally collected data show similar quality TPMs to those included in our study and show areas of predicted tumor beyond contrast enhancement. External Data 3 shows additional histology from biopsy-confirmed tumor in a non-enhancing area of predicted high tumor probability pre-surgery. B) Examples of pTOC and no pTOC, showing patients with predicted non-enhancing tumor presence compared to patients with no predicted tumor presence outside of contrast enhancement. C) Kaplan-Meier survival curves showing longer overall survival in patients with pTOC pre-surgery (HR = 1.67 (1.07-2.62), p = 0.027), similar to the pattern observed with autopsy-confirmed tumor outside contrast prior to death seen in Figure 2.

## 4. Discussion

This study developed a multi-stage predictive model that identifies areas of tumor beyond traditional imaging signatures. A pathological tumor prediction model based on histology segmentations was combined with MRI-based maps of tissue composition to bridge the gap between histopathological assessment and non-invasive glioma tracking. We validated this technique using autopsy tissue samples as ground truth, and successfully applied our models to external data to demonstrate generalizability. This study is the first of its kind to provide tumor probability maps validated using tissue well-beyond the current treatment margin, and is, to date, the largest study of radio-pathomic signatures at autopsy in brain cancer.

This study demonstrates that around half of gliomas exist in part beyond the treated margin, with higher rates observed in higher grade tumors such as glioblastomas. Since this study only used tissue samples from areas of suspected tumor that could be aligned accurately to the MRI after tissue processing, this frequency is likely a lower bound, as it is possible that areas of tumor are missed in the tissue sample selection process. Future targeted MRI-based research should focus on these regions specifically to improve non-invasive tumor localization. This is particularly crucial to understanding tumor in the treated state, as our preliminary assessment of treatment effects on TOC highlight treatment’s impact on radio-pathomic relationships. Bevacizumab in combination with Rad+TMZ treatment was associated with greater TOC compared to bevacizumab-naïve patients, which is hypothesized to be caused by the anti-angiogenic treatment disrupting the co-occurrence of tumor and contrast enhancement. In the future, targeted research should be conducted to elucidate the mechanism of the relationship between treatment and imaging signatures.

To detect regions of tumor beyond traditional imaging signatures, we developed a multistage model to predict tumor using autopsy tissue samples as ground truth. Prior research attempting similar tasks have used biopsy tissue samples which inherently fail to capture tumor beyond the treated margin^18,31–33^. Our prior studies of autopsy tissue samples highlighted how imaging signatures validated within contrast enhancement, such as an inverse relationship between ADC and cellularity, show much weaker relationships when validated using tissue from beyond the enhancing region^14,16^. In addition to providing TPMs, the intermediate tissue segmentations for cellularity, ECF, and cytoplasm aided in interpreting different tumor features, such as distinguishing between areas of PN and non-PN tumor, as well as differentiating between tumor and vasogenic edema within FLAIR hyperintensity. Additionally, using post-treatment pathology aligned to MR images acquired post-treatment allows for the model to differentiate treatment-related effects from areas of active tumor, underscoring the potential utility for TPMs as a tumor monitoring tool in later clinical stages of the disease. This highlights how providing a rich pathological ground truth can vastly improve non-invasive tumor tracking, particularly using machine learning techniques to map relationships invisible to the naked eye.

We additionally applied our model to an independent dataset to assess predicted tumor presence prior to surgery. Predicted TOC was observed in 69 percent of cases and showed worse survival rates compared to patients with well-circumscribed tumors. This suggests that TPMs identify areas of tumor pre-treatment, which may aid in stratifying patient risk in early stages of the disease. The frequency of predicted areas of non-enhancing tumor may provide a more accurate estimate of TOC frequency, as the TPMs are able to assess tumor presence across the entire brain instead of select tissue samples. Though future research using biopsy samples from a greater number of patients is required to confirm tumor presence at timepoints prior to death, these results suggest that TPMs may identify tumor prior to its visual appearance on traditional imaging. By applying this technique to retrospective data and previously collected clinical trials, these maps may be able to reveal new signatures of treatment response in terms of reduced hypercellular volume and low tumor probability, which in turn could identify subsets of patients that selectively respond to treatment.

### 4.1. Limitations

Due to the use of autopsy tissue samples, the time between imaging and death is an important limiting factor in this study. Tumor likely continues to grow near death, and imaging acquired months before death may not reflect how the patient’s imaging would look immediately prior to death. We have accounted for this in our study by restricting our frequency assessment of TOC to imaging within two months of death and have matched our train and test data sets for time between MRI and death. Future studies using longitudinal imaging to model tumor growth rates may control for this factor more precisely. Potential tissue distortions from processing may result in imperfect alignment with MRI data; however, our previously published tissue processing protocol controls for this at every step of tissue collection, and regions of interest defined after warping are used to exclude regions of suboptimal tissue quality. Due to differences in background values across subjects and presence of small, localized regions of high tumor probability amongst areas of low tumor probability, we defined tumor presence as an increase in tumor probability relative to surrounding tissue and contralateral regions. Future work examining TPMs across specific tumor types in larger samples may be able to define specific cutoffs, but this remains an area of active work. Additionally, while this is the largest study of its kind using an unprecedented amount of autopsy tissue data aligned to MRI, the validation set for this study was relatively small compared to other machine learning studies. Our predictions on external data sets highlight the feasibility of applying this method to large data sets for further validation, thus precise validation using biopsy tissue is being conducted as a follow-up to this study.

### 4.2. Conclusion

Autopsy tissue samples aligned to MRI were used to develop a multi-stage model for tumor probability to predict areas of non-enhancing tumor that occur in glioma patients. This technique has the potential to improve disease tracking in the post-treated state and could expand the treated margin to encompass additional infiltrative tumor missed by current tracking techniques.

## Data Availability

The MRI and histology data sets used in this study are not currently publicly available due to data usage agreement restrictions. De-identified data is available upon reasonable request.

## 7. Supplemental Material

**Table 1:** Demographic and clinical summary for included subjects)

|  | Train | Test |
| --- | --- | --- |
| **Number of Subjects** | 43 | 22 |
| **Age (years)** | 58.4 (14.1) | 60.9 (11.7) |
| **Overall Survival (months)** | 48.8 (71.2) | 34.8 (42.3) |
| **GBM/Non-GBM** | 29 / 14 | 18 / 4 |
| **Radiation treatment (y/n)** | 39 / 4 | 21 / 1 |
| **Chemotherapy (y/n)** | 39 / 4 | 20 / 2 |
| **Bevacizumab (y/n)** | 34 / 9 | 15 / 7 |
| **Tumor Treating Fields (y/n)** | 17 / 26 | 6 / 16 |

**Supplemental Figure 1:**
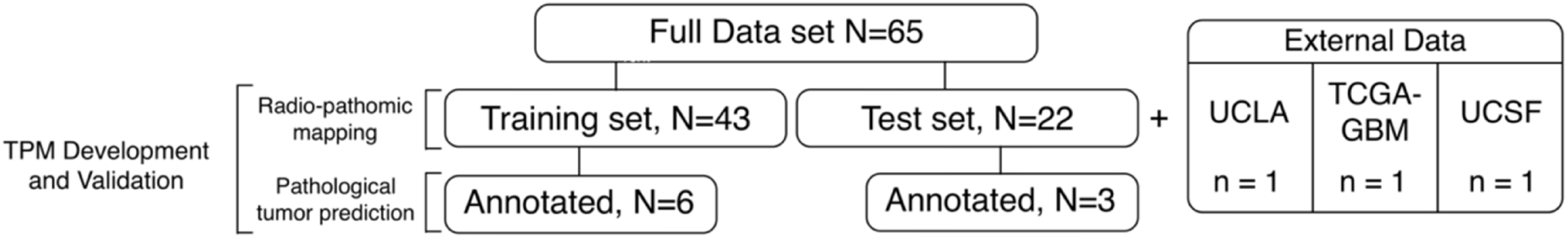
Flowchart for the subjects included in each stage of the tumor probability map (TPM) model development

